# Association of electronic cigarette use with chronic kidney disease in NHANES 2017–2020: A replication study

**DOI:** 10.1101/2025.10.09.25337422

**Authors:** Gal Cohen, Sooyong Kim, Arielle Selya

## Abstract

Combustible cigarette (CC) smoking is a modifiable risk factor for chronic kidney disease (CKD), but the CKD risk associated with electronic cigarette (EC) use, relative to CC use or nonuse, is unclear. We read with interest a recent analysis (Li et al., 2025) of the NHANES 2017–2020 database that evaluated this research question. In the course of verifying the results, we identified multiple concerns in the reported methodology including a) not *a priori* segmenting into CC current, former, and never use to minimize confounding between CC and EC use in subsequent analysis; b) unclear rationale for excluding ~80% of the available sample; and c) performing a sensitivity analysis on an EC cohort with n=1 harm events. The present replication study indicates that, after segmenting for CC use history and adjusting for covariates, particularly former smoking, there was no association between EC current use (past 5 days) and CKD risk in this NHANES population. This replication study highlights how precise segmentation and characterization of exposure to all tobacco products is required for accurate analysis of cross-sectional studies and suggests metrics for incorporation into future waves of NHANES.

## Introduction

Chronic kidney disease is the 8th leading cause of mortality in the U.S., with a prevalence of over 35 million adults (1). Risk factors include age, diabetes, and hypertension. Combustible cigarette (CC) smoking is a known modifiable risk factor; consequently, evaluating the association of electronic cigarette (EC) use with this disease is relevant. We read with interest a recent analysis (Li et al., 2025), which comprised a retrospective cross-sectional analysis of the NHANES 2017–2020 database to evaluate this research question (2). The authors reported that current EC use was associated with an increased incidence of CKD and that CKD risk was also positively correlated with the frequency of EC use. As described below, multiple aspects of the analysis appeared to not conform to best practices for optimizing precision and accuracy (3). Therefore, a replication study was performed to verify the findings.

## Methods

Data were downloaded from the NHANES “Pre-Pandemic” dataset spanning 2017– 2020. The separate 2017–2018 and 2019–2020 files were not used, as they are already incorporated into the 2017–2020 dataset. These data are available at https://wwwn.cdc.gov/nchs/nhanes/continuousnhanes/default.aspx?Cycle=2017-2020 (accessed 9/5/2025).

### Definitions

1. EC current use was assessed as use in the past 5 days (SMQ690H, SMQ849). In 2019, the NHANES discontinued asking about EC ever-use and EC past 30-day use (SMQ900, SMQ905), so this was the only metric available for the complete 2017–2020 dataset (see the Discussion for suggestions for future NHANES waves).
2. Current established CC use was derived from two variables: (a) smoking 100+ cigarettes/lifetime and (b) currently smoking cigarettes “every day” or “some days” (vs. not at all”).
3. CC former established use was derived from two variables: (a) smoked 100+ cigarettes/lifetime and (b) now smoking “not at all” (vs. “every day” or “some days”).
4. CC never established use was defined as having smoked <100 cigarettes/lifetime.
5. The glomerular filtration rate (GFR) was calculated via the 2021 CKD-EPI Creatinine Equation, which estimates the glomerular filtration rate (eGFR) using age, sex, and serum creatinine as variables. eGFR (mL/min/1.73 m^2^) = 142 * min(SCr/κ, 1)^α^ * max(SCr/κ, 1)^-1.200^ * 0.9938^AGE^ * 1.012[if female] where SCr (serum creatinine) in mg/dL (LBXSCR in NHANES) k = 0.7 (females) or 0.9 (males) α = −0.241 (females) or −0.302 (males) AGE in years
6. CDK metrics (see Figure 1)
  A) The KDIGO CKD Heat Map provides the most precise assessment of chronic kidney disease (CKD) progression, morbidity, and mortality risk (4). The columns represent the albumin/creatinine ratios, and the rows represent the estimated glomerular filtration rates. The color coding represents the risk of progression to CKD. Five gradations of risk exist: low risk of progression to CKD, moderate risk, high risk, very high risk and highest risk. Note that a definitive assessment of CKD requires the presence of symptoms for at least 3 months and a clinical evaluation in addition to the Heat Map metrics.
  B) Li et al. utilized two definitions of CKD, which had lower precision (binary scales representing a positive or negative result). In the narrow definition, an albumin/creatinine ratio of 300+ represents a positive result.
  C) In the broader definition of Li et al., an albumin/creatinine ratio of 30+ or an eGFR < 60 represents a positive result.

**Figure 1.**
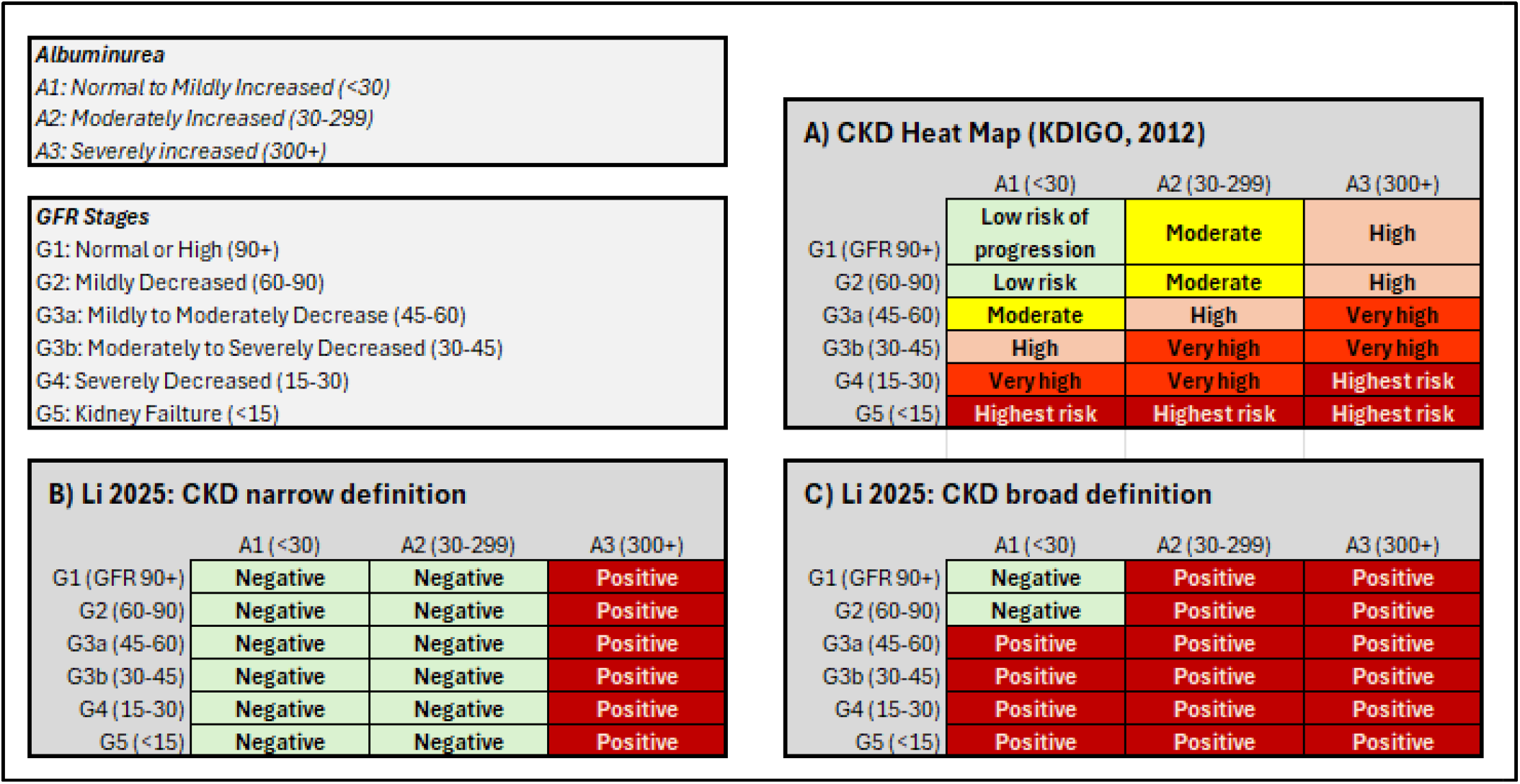
CKD assessment scales. A. KDIGO CKD heatmap. The columns represent the albumin/creatinine ratio, and the rows represent the estimated glomerular filtration rate. The color coding represents the risk of CKD progression, morbidity, and mortality (green represents low risk, yellow represents moderate risk, and red represents high risk). B. CKD narrow definition from Li et al., 2025. An albumin/creatinine ratio of 300+ represents a positive result. C. CKD broad definition from Li et al., 2025. An albumin/creatinine ratio of 30+ or an eGFR < 60 represents a positive result. CKD (Chronic Kidney Disease); KDIGO (Kidney Disease: Improving Global Outcomes)

## RESULTS

### I. Analytic sample

An attempt was made to replicate the analysis dataset of the authors. As shown in Figure 2, from a starting sample of N=9,693 adults, the retained analytic sample of N=8,109 consisted of 203 EC-current-users and 7,906 non-EC current-users after the exclusions were enumerated owing to missing data. The missing data for the 1,584 excluded individuals primarily pertained to EC use status, albumin/creatinine levels, and metrics used to calculate the estimated GFR.

**Figure 2.**
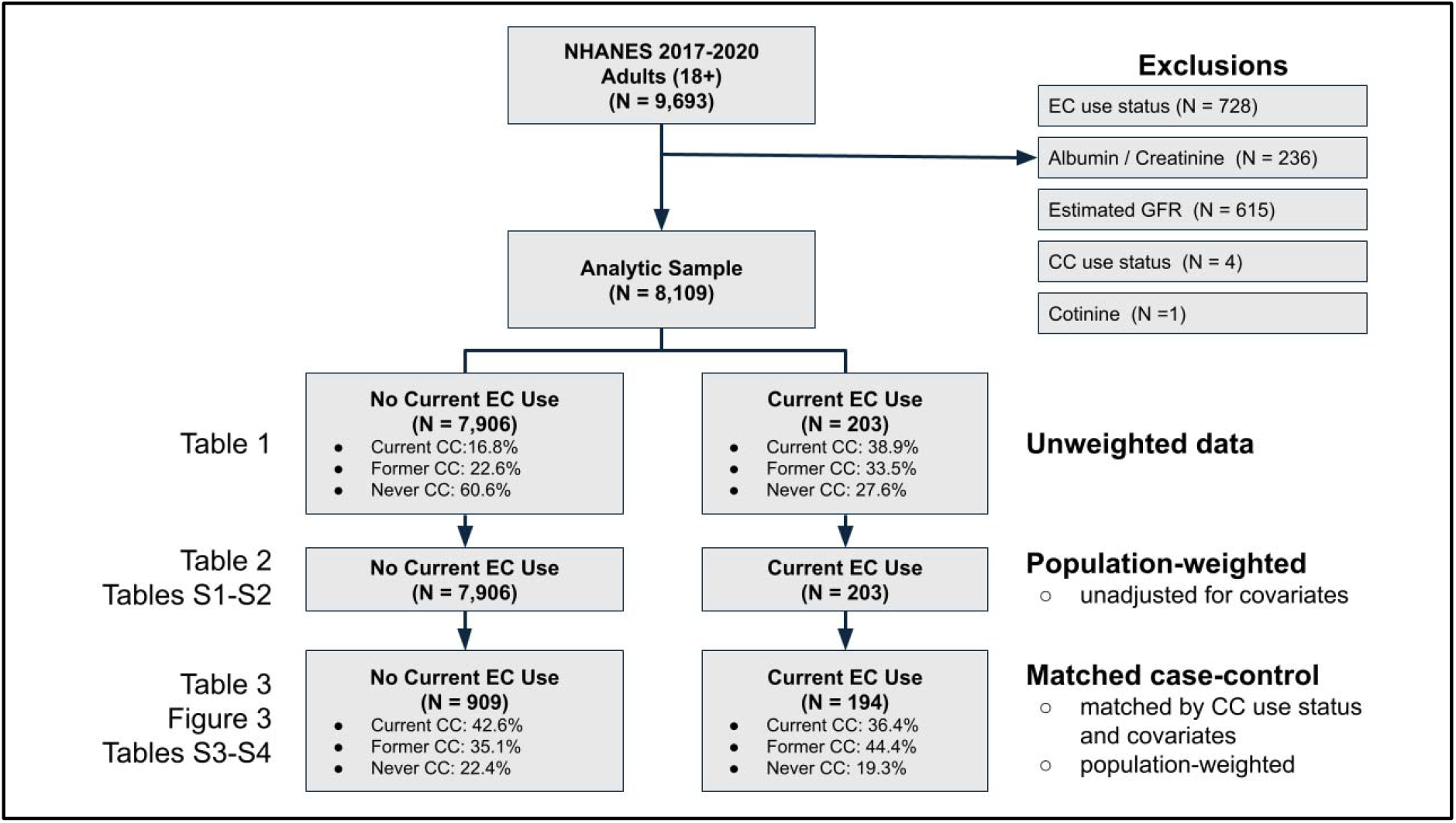
Analytic sample, NHANES 2017–2020.

### II. Unweighted data (segmented by CC and EC use)

NHANES unweighted data (i.e., raw data) are provided in Table 1. Consistent with best practices, the sample was segmented by CC and EC use histories to minimize confounding due to the nonindependence of CC and EC use patterns (3).

**Table 1.**
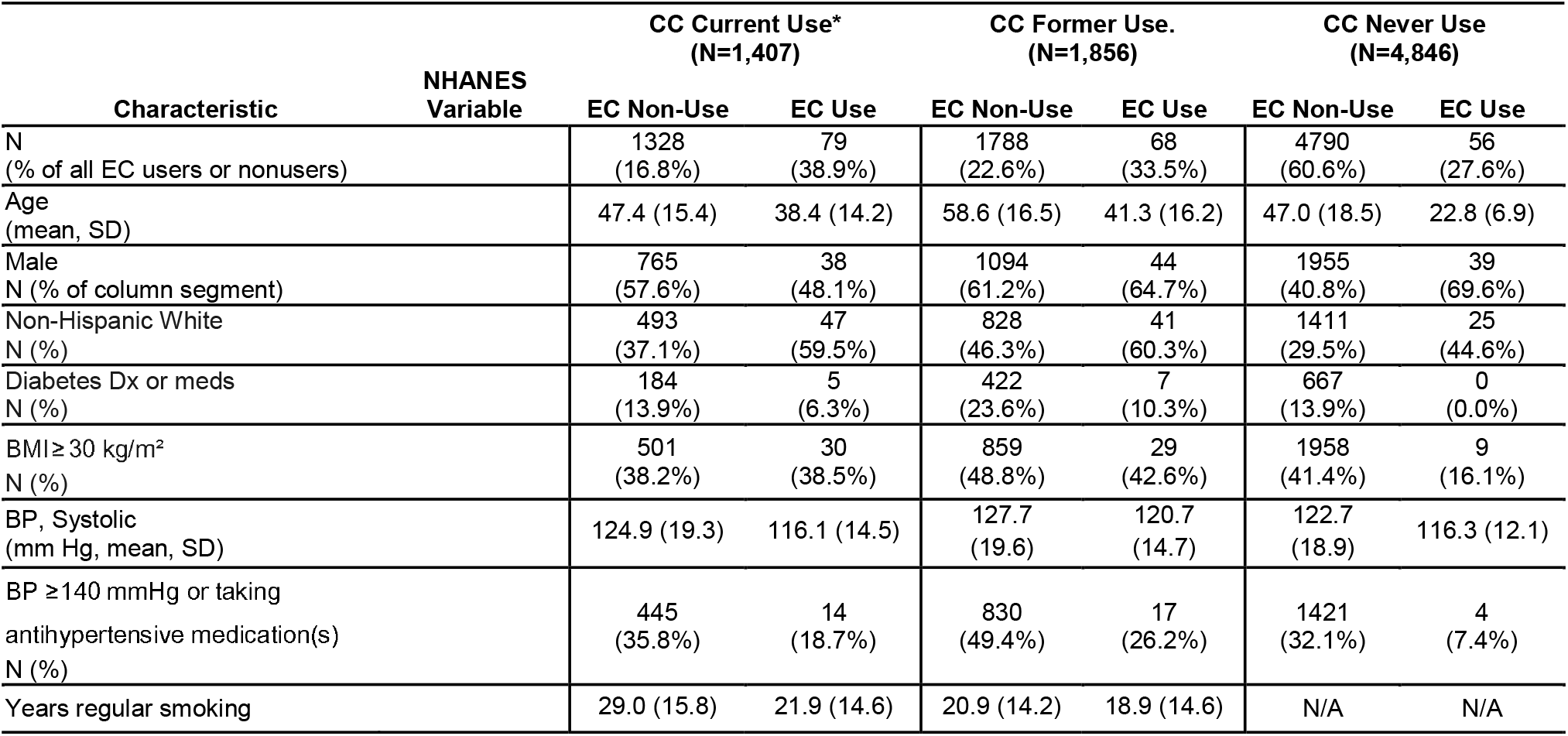
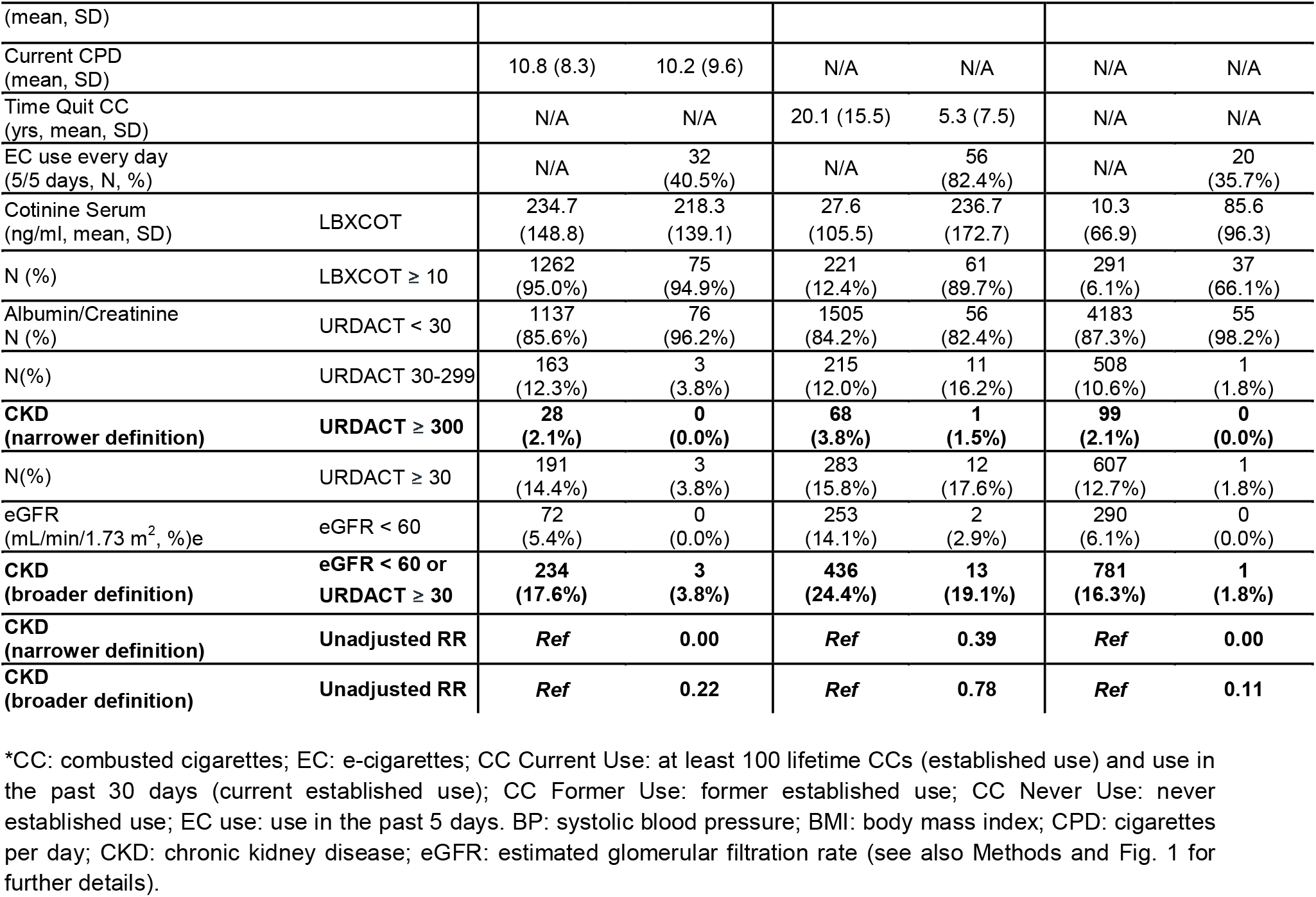
Unweighted analysis sample (segmented by CC and EC use)

In the unweighted data, large between-group imbalances existed in the demographic covariates associated with CKD risk. The EC groups had lower rates of diabetes, hypertension and obesity than did the non-EC groups. Segmentation by CC use history revealed additional segment-specific differences. EC users who were previously CC users had stopped smoking much more recently than former CC users who were not EC users (5.3 vs. 20.1 years quit, respectively). Those individuals who had completely switched from CC use to EC use also had much higher rates of daily EC use than did those who were CC dual-users or never-users (82.4% vs. 40.5% and 35.7%, respectively) and were older than CC never-users were (41.3 years old vs. 22.8 years old). Thus, daily EC use was most often observed in cohorts at higher risk for CKD due to non-EC covariates (older, long-term CC users who had quit in recent years).

Li et al. utilized two definitions of CKD, both of which are reported in Table 1. A broader definition included individuals with a glomerular filtration rate (GFR) < 60 or an albumin/creatinine ratio ≥ 30. A narrower definition was limited to individuals with an albumin/creatinine ratio ≥ 300. In the raw data, the incidence of CKD, according to the broader definition, was markedly lower in each EC-using group (vs. the comparator EC nonusing group): current smokers (3.8% vs. 17.6%), former smokers (19.1% vs. 24.4%), and never smokers (1.8% vs. 16.3%). Using this broader definition of CKD, the unadjusted RR for EC users vs. nonusers was 0.22 for CC current users, 0.78 for CC former users, and 0.11 for CC never users. The lower incidence of CKD held true for the narrower definition of CKD as well (0.5% for EC users vs. 2.5% for EC nonusers; RR of 0.2).

The higher rates of CKD-related endpoints in the EC nonusers than in the EC-users were consistent with imbalances in non-EC covariates that are known risk factors for CKD, such as age, diabetes, and hypertension. Additionally, there was a higher rate of CKD-related endpoints in CC former-users, and this was true for both EC nonusers and EC users.

Critically, only 1 EC user met the narrower definition of CKD. A sensitivity analysis was performed by Li et al., despite the number of harm events being below the threshold for a valid statistical analysis. Seventeen EC users met the broader CKD definition of Li et al., of which 13 formerly smoked and 3 were dual-users.

### III. Population-weighted data

In the population-weighted data that account for NHNAES oversampling of racial/ethnic minorities, the increased prevalence of non-Hispanic white individuals was narrowed but still persisted for EC users who were CC current and former users (see Table S1). In the EC groups, age remained younger, male sex remained more common, and rates of diabetes, hypertension and obesity remained lower than those in the non-EC cohorts. The former CC users had stopped smoking 18.7 years previously in the non-EC group, whereas the EC group had stopped CC use more recently (4.0 ± 5.6 years, indicating many recent CC users).

In the population-weighted data, segmented by CC use, the OR of CKD (using the broader definition of Li et al.) was lower in each of the EC use groups than in the comparable nonuse groups (Table 2). Critically, while p values can be derived, the low number of harm events limits the statistical reliability of the analysis.

**Table 2.** Univariate associations between current ENDS use and CKD (NHANES 2017– 2020 unadjusted, population-weighted data).

| Characteristic | OR (CI)<br>(EC use vs. nonuse) | P value | Number of EC CKD events<br>in in raw data sample<br>(broader definition of Li et al., 2025) |
| --- | --- | --- | --- |
| CC current-users | 0.49 (0.12–2.05) | 0.312 | n=3 |
| CC former-users | 0.82 (0.33–2.06) | 0.659 | n=13 |
| CC never-users | 0.02 (0.00–0.18) | 0.001 | n=1 |

### IV. Matched case–control samples

Current-, former-, and never-smoking EC users were matched to EC nonusers on the basis of covariates with substantial imbalance (Table 1) to comprise a combined case–control sample. A total of 194 EC users were matched to 909 EC nonusers, and their characteristics are summarized in Table 3.

**Table 3.** Matched, covariate-adjusted and population-weighted analysis samples.

| Characteristic | Comment | Matched Case-Control |  |
| --- | --- | --- | --- |
|  |  | EC Non-Use | EC Use |
| N | Unweighted | 909 | 194 |
| Age (mean, SD) | Weighted mean (SD) | 35.8 (14.3) | 34.2 (14.4) |
| Male (%) | Weighted % | 59.3% | 62.8% |
| Non-Hispanic White (%) | Weighted % | 70.1% | 75.9% |
| Diabetes Dx or meds (%) | Weighted % | 6.8% | 4.6% |
| BMI $\geq 30$ kg/m<sup>2</sup> (%) | Weighted % | 37.0% | 32.2% |
| BP, Systolic (mm Hg, mean/SD) | Weighted mean (SD) | 117.2 (13.6) | 118.3 (13.6) |
| BP $\geq 140$ mmHg or taking antihypertensive medication(s) (%) | Weighted % | 18.7% | 18.7% |
| Current CC use (%) | Weighted % | 42.6% | 36.4% |
| Former CC use (%) | Weighted % | 35.1% | 44.4% |
| Never CC use (%) | Weighted % | 22.4% | 19.3% |
| Yrs. regular smoking (mean, in current and former CC users) | Weighted mean (SD) | 19.8 (13.9) | 18.3 (14.0) |
| Current CPD<br>(mean, in current CC users) | Weighted % | 12.7 (8.6) | 11.2 (9.8) |
| Time Quit CC<br>(yrs, mean, in former CC users) | Weighted % | 5.5 (7.0) | 4.0 (5.6) |
| EC use every day (5/5 days, %) | Weighted mean (SD) | N/A | 63.9% |
| Cotinine Serum (ng/ml, mean) | LBXCOT | 110.9 (155.4) | 211.8 (151.8) |
| Cotinine $\geq 10$ ng/ml N (%) | LBXCOT $\geq 10$ | 47.7% | 88.5% |
| Albumin/Creatinine N (%) | URDACT $< 30$ | 93.1% | 91.0% |
|  | URDACT 30-299 | 5.9% | 8.8% |
| **CKD (narrower definition)** | **URDACT $\geq 300$** | **1.0%** | **0.2%** |
| | URDACT $\geq 30$ | 6.9% | 9.0% |
| eGFR (mL/min/1.73 m<sup>2</sup>, %) | eGFR $< 60$ | 0.9% | 0.7% |
| **CKD (broader definition)** | **eGFR $< 60$ or URDACT $\geq 30$** | **7.4%** | **9.3%** |
| **CKD (narrower definition)** | **RR\*** | **Ref** | **0.20\*** |
| **CKD (broader definition)** | **RR\*** | **Ref** | **1.26\*** |

Note: estimates refer to weighted % for categorical variables and weighted mean (weighted SD) for continuous variables. \*RR (relative risk) calculated based on the prevalence of CKD among EC users divided by the prevalence of CKD among EC nonusers. No formal modeling analysis was conducted due to the limited sample size.

In the matched controls, according to the definitions of Li et al., the RR for CKD was lower for the narrower definition (0.17) but higher for the broader definition (1.26) for EC users than for EC nonusers. According to the more precise KDIGO heatmap definition risk of CKD progression, morbidity or mortality, there was no apparent difference between EC users and nonusers. In the EC use cohort, 90.7% were considered low risk, 8.7% were moderate risk and 0.6% were high risk. In the EC nonuse cohort, 92.6% were considered low risk, 5.9% were moderate risk and 1.5% were high risk (see Figure 3).

**Figure 3.**
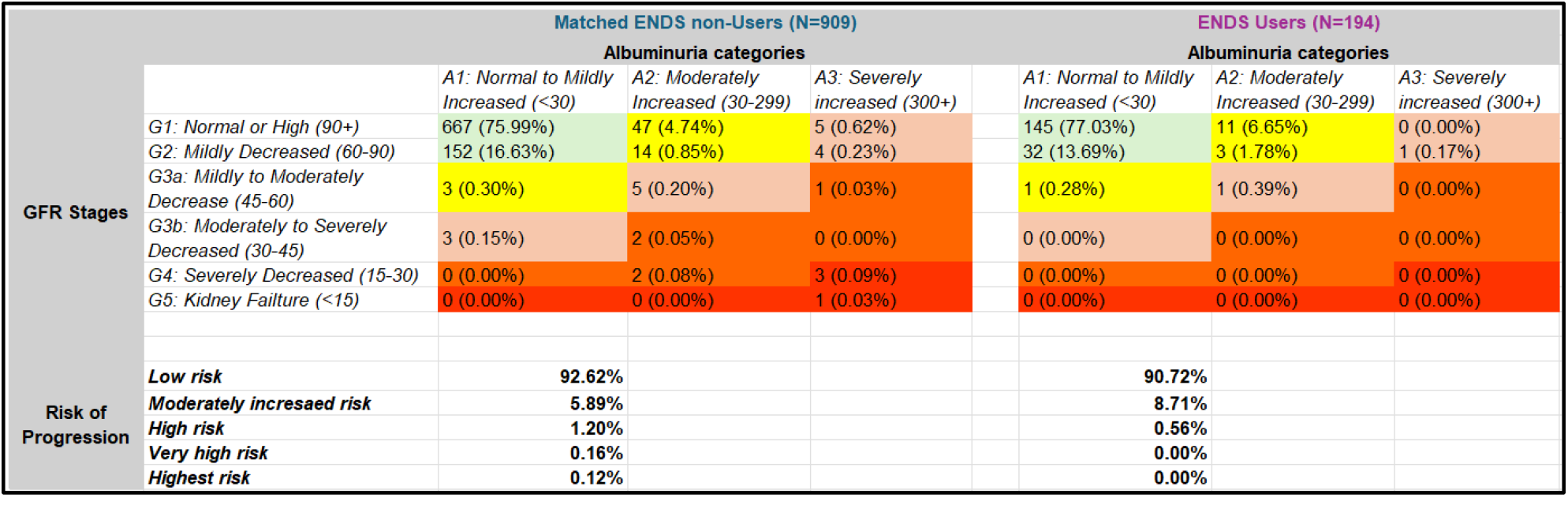
Risk of progression to CKD (matched, covariate-adjusted and population-weighted analysis samples). Risk of progression per the CKD Heat Map (4).

## Discussion

Cross-sectional studies compare samples that are by definition unbalanced and then use modeling approaches to account for any significantly impact covariates except for the variable of interest. In the case of cross-sectional studies of nicotine products, because the use patterns of noncombusting nicotine products are not independent of each other, precise segmentation and characterization of exposure can mitigate confounding and improve accuracy (3).

We note imprecision and deviations from best practices in the analysis of Li et al:

1. Descriptive data for CKD prevalence between groups were not published by the authors. While the underlying dataset is available from NHANES, the number of calculations and exclusions needed to derive the analysis data indicate that a new dataset was generated. It is central in every cross-sectional study to be able to transparently follow how the raw data were transformed into adjusted odds ratios.
2. The present analysis resulted in a retained analytic sample size of n = 7,139, which was much larger than Li et al.’s sample size of n = 872 (n = 684 EC nonusers and n = 188 EC users). One potential explanation for this discrepancy might be failure to account for skip-patterns in the survey: some questions are asked in series, where a second question is asked depending on the answer of the first question. Non answer to the second question (e.g., current smoking status) is not the same as missing or incomplete data if it is answered in the first question (i.e., never-established smoking).
3. Li et al. reported that granular data were not available about CC use history and consequently omitted critical variables, including years of CC use, former CC use, time quit and CPD; however, these data are readily available (and were included in the present analysis).
4. Because smoking and vaping use patterns are not independent, it is crucial to segment current, former, and never smokers separately in the EC group (and comparatively in the non-EC group) to minimize confounding. As lingering health effects from prior CC use could be misattributed to EC use, EC users who have switched from former smoking should most precisely be compared with former smokers with a similar duration of CC smoking and time quit. Likewise, EC users who dual-use CCs should be compared with exclusive CC smokers who have a similar duration of CC smoking (3).
5. Li et al. used imprecise definitions of CKD to obtain binary (yes/no) metrics (see Methods). In the sensitivity analysis of Li et al. involving the more stringent definition of CDK, the EC group had only n=1 harm event (URDACT 300+), rendering the analysis invalid. As a statistical best practice, cross-sectional studies should include only cohorts with a ratio of at least 10 harm events per adjustment variable in the regression model.(3) Furthermore, the CKD Heat Map provides more precise gradations, and represents risk more accurately, such as the risk of progression to CKD, rather than a definitive diagnosis.(4) In addition, the diagnosis of CKD requires sustained aberrant readings for 3 months to exclude transient albuminuria associated with events unrelated to CKD (e.g., infection, fever, vigorous exercise) or acute kidney injury that may be reversed in the near future.
6. Li et al. reported a dose–response relationship between the frequency of EC use and CKD risk. However, this is likely explained by confounding, not a true dose–response effect, as frequent EC use was concentrated within *formerly-* smoking EC users (54/65 = 83%) as opposed to EC users who currently (32/79 = 41%) or never smoked (17/50 = 34%). These former smokers were older and more likely to have comorbidities (Table 1).
7. Finally, CKD is, by definition, a chronic condition. Without information about the timing of CKD onset vs. the duration of EC use, establishing causality is not possible. In particular, the timing of the onset of CKD and other comorbidities is necessary to identify people who may have switched from CCs to ECs *because* they were experiencing smoking-related health effects (reverse-causality) and to perform a sensitivity analysis on the inclusion of this population.

This replication study also highlights the importance of transparency and comprehensiveness of the disclosure of adjusted and unadjusted data. Complete disclosure in the text and supplemental information provides visibility into the exact methods used by the authors and any strengths and limitations of the analysis. For example, the first step of converting raw data into population-weighted data is a common source of error. Authors from Research Update have recently published a series of four analyses of this NHANES database with impossible sample sizes, of which only one has been corrected (via retraction by an editorial board) as of October 2025 (5–9). However, a highly cited meta-analysis revealed that one of its primary results was brittle with respect to one of these papers, and this major error also remains uncorrected (3,10).

## Conclusion

In contrast to the study of Li et al., this replication study did not find a relationship between EC use and CKD. This was likely due to the imprecision in the analytic methods of Li et al. Lack of transparency and completeness of disclosure of adjusted and unadjusted datasets likely also limited the efficacy of the peer review process to identify and correct these imprecisions.

This replication study was subject to several limitations. First, chronic kidney disease diagnosis requires persistent aberrant eGFR or urine albumin readings for 3 months to rule out acute conditions such as nonkidney infections, but the endpoints here were limited to a single acute timepoint (11). Next, the only EC use measure available in recent NHANES is past 5-day use, without duration of use, or 30-day use, so dose–response analyses are best addressed via PATH or other databases which are more detailed than NHANES. The lack of tracking duration of EC use means that there is no means to determine which came first, the harm event, or EC use. Both the time of diagnosis and the time of starting regular EC use are necessary to help establish causality. Finally, there was a minimal number of harm events in the EC group, meaning that sensitivity analyses were not valid in this dataset.

In summary, this replication study is instructive because it provides an example of common issues that should be taken into account for more precise and accurate design and analysis of cross-sectional studies of nicotine and tobacco use. This case study may also be instructive for peer reviewers assessing studies for publication. Crucially, this study highlights the need for NHANES to include more comprehensive nicotine product use metrics (including duration of use and past 30-day use) in future waves to improve the precision and accuracy of subsequent analyses. The important question of whether nicotine product use induces and/or reduces the risk of kidney injury or other diseases should be studied via more precise and accurate methodologies.

## Data Availability

All data produced in the present study are available upon reasonable request to the authors. Source data is available at https://wwwn.cdc.gov/nchs/nhanes/continuousnhanes/default.aspx?Cycle=2017-2020

## Authors’ contributions

G.C. wrote the initial draft. All the authors contributed to the writing, editing and review of the manuscript. S.K. was the lead for the analysis of NHANES data. All the authors contributed to the review of the data analysis. All the authors have read and approved the manuscript.

