## Supplement I for "Association of electronic cigarette use with chronic kidney disease in NHANES 2017–2020: A replication study"

**Supplemental information**

**Table S1** NHANES 2017–2020 population-weighted data, unadjusted for covariates

|  |  | **CC Current Use***  **(N=1,407)** | | **CC Former Use.**  **(N=1,856)** | | **CC Never Use**  **(N=4,846)** | |
| --- | --- | --- | --- | --- | --- | --- | --- |
| **Characteristic** | **NHANES**  **Variable** | **EC Non-Use** | **EC Use** | **EC Non-Use** | **EC Use** | **EC Non-Use** | **EC Use** |
| N |  | 1328 | 79 | 1788 | 68 | 4790 | 56 |
| Age (mean, SD) |  | 44.5 (15.1) | 39.3 (13.7) | 54.6 (16.1) | 35.2 (14.9) | 45.9 (17.9) | 23.0 (6.1) |
| Male (%) |  | 53.4% | 51.7% | 58.1% | 71.2% | 42.2% | 70.8% |
| Non-Hispanic White (%) |  | 62.2% | 79.5% | 72.2% | 80.7% | 58.4% | 59.1% |
| Diabetes Dx or meds (%) |  | 10.6% | 3.6% | 17.7% | 6.8% | 10.6% | 0.0% |
| BMI ≥ 30 kg/m² |  | 35.5% | 34.4% | 47.7% | 36.3% | 41.5% | 12.7% |
| BP, Systolic (mm Hg, mean) |  | 122.2 | 116.8 | 124.8 | 120.7 | 120.3 | 115.5 |
| BP ≥140 mmHg or taking antihypertensive medication(s)  (%) |  | 29.8% | 19.3% | 39.3% | 24.5% | 26.8% | 4.5% |
| Yrs. regular smoking (mean. SD) |  | 26.9 (15.1) | 23.5 (14.0) | 19.0 (13.4) | 14.2 (12.6) | N/A | N/A |
| Current CPD (mean, SD) |  | 11.9 (8.3) | 10.8 (9.5) | N/A | N/A | N/A | N/A |
| Time Quit CC (yrs, mean) |  | N/A | N/A | 18.7 (15.1) | 4.0 (5.6) | N/A | N/A |
| EC use every day (5/5 days, %) |  | N/A | 46.2% | N/A | 84.5% | N/A | 48.0% |
| Cotinine Serum (ng/ml, mean) | LBXCOT | 226.8 (148.2) | 219.2 (127.0) | 26.8 (103.2) | 247.7 (162.7) | 7.8 (55.5) | 100.0 (104.5) |
| N (%) | LBXCOT **≥** 10 | 93.8% | 96.6% | 12.7% | 90.6% | 4.7% | 68.5% |
| Albumin/Creatinine  N (%) | URDACT < 30 | 90.2% | 93.8% | 88.8% | 85.9% | 90.1% | 99.7% |
|  | URDACT 30-299 | 7.9% | 6.2% | 9.0% | 13.7% | 8.5% | 0.3% |
| **CKD**  **(narrower definition)** | **URDACT ≥ 300** | **1.9%** | **0.0%** | **2.2%** | **0.4%** | **1.4%** | **0.0%** |
|  | URDACT **≥** 30 | **9.8%** | **6.2%** | **11.2%** | **14.1%** | **9.9%** | **0.3%** |
| eGFR  (mL/min/1.73 m^2^, %) | eGFR < 60 | **3.2%** | **0.0%** | **9.2%** | **1.5%** | **4.7%** | **0.0%** |
| **CKD**  **(broader definition)** | **eGFR < 60**  **or URDACT ≥ 30** | **12.0%** | **6.2%** | **17.4%** | **14.7%** | **13.0%** | **0.3%** |
| **CKD**  **(narrower definition)** | **Unadjusted RR** | ***Ref*** | **0.00** | ***Ref*** | **0.18** | ***Ref*** | **0.00** |
| **CKD**  **(broader definition)** | **Unadjusted RR** | ***Ref*** | **0.52** | ***Ref*** | **0.84** | ***Ref*** | **0.02** |

CC: combusted cigarettes; EC: e-cigarettes; CC Current Use: at least 100 lifetime CCs (established use) and use in the past 30 days (current established use); CC Former Use: former established use; CC Never Use: never established use; EC use: use in the past 5 days. BP: systolic blood pressure; BMI: body mass index; CPD: cigarettes per day; CKD: chronic kidney disease; eGFR: estimated glomerular filtration rate. See Methods and Fig. S1 for further details.

**Table S2** Replication of Li et al., using non case‒control sample, unbalanced for CC current and former use history

|  | **Adjusted for** | **OR (CI)** | **P value** |
| --- | --- | --- | --- |
| **Model 1** | Age, gender, race | 1.32 (0.56–3.13) | 0.508 |
| **Model 2** | Model 1 + BMI | 1.33 (0.56–3.19) | 0.497 |
| **Model 3** | Model 2 + diabetes, hypertension, current smoking | 1.40 (0.58–3.41) | 0.431 |

(Age, BMI: continuous; diabetes, hypertension: binary; current smoking: modified the smoking status variable into current vs. former or never)

**Table S3** Matched, covariate-adjusted and population-weighted samples (segmented by CC and EC use)

| **Characteristic** | **Comment** | **CC Current Use**  **(N=446)** | | **CC Former Use.**  **(N=365)** | | **CC Never Use**  **(N=292)** | |
| --- | --- | --- | --- | --- | --- | --- | --- |
|  |  | **EC Non-Use** | **EC Use** | **EC Non-Use** | **EC Use** | **EC Non-Use** | **EC Use** |
| N |  | 371 | 75 | 300 | 65 | 238 | 54 |
| Age  (mean, SD) | Weighted mean (SD) | 39.0 (13.7) | 38.6 (13.7) | 39.8 (14.2) | 35.4 (15.0) | 23.5 (6.8) | 23.1 (6.2) |
| Male  (%) | Unweighted N (Weighted %) | 215 (52.2%) | 36 (49.8%) | 179 (60.7%) | 41 (70.4%) | 156 (70.6%) | 37 (69.6%) |
| Non-Hispanic White  (%) | Unweighted N (Weighted %) | 218 (76.2%) | 44 (77.8%) | 115 (65.3%) | 39 (80.7%) | 109 (66.0%) | 25 (61.4%) |
| Diabetes Dx or meds  (%) | Unweighted N (Weighted %) | 25 (5.8%) | 5 (4.0%) | 43 (12.2%) | 7 (7.0%) | 0 (0.0%) | 0 (0.0%) |
| BMI ≥ 30 kg/m²  (%) | Unweighted N (Weighted %) | 134 (35.6%) | 30 (38.0%) | 150 (51.4%) | 28 (35.8%) | 44 (17.0%) | 9 (13.2%) |
| BP, Systolic  (mm Hg, mean) | Weighted mean (SD) | 117.8 (13.7) | 116.8 (14.1) | 119.2 (14.1) | 120.7 (14.1) | 113.2 (12.1) | 115.5 (10.6) |
| BP ≥140 mmHg or taking antihypertensive medication(s)  (%) | Unweighted N (Weighted %) | 83 (22.1%) | 14 (19.3%) | 92 (23.7%) | 17 (24.5%) | 5 (4.3%) | 4 (4.5%) |
| Yrs. regular smoking  (mean) | Unweighted N (Weighted %) | 22.2 (14.4) | 23.1 (14.2) | 16.9 (12.6) | 14.4 (12.6) | N/A | N/A |
| Current CPD  (mean) | Unweighted N (Weighted %) | 12.7 (8.6) | 11.2 (9.8) | N/A | N/A | N/A | N/A |
| Time Quit CC  (yrs, mean) | Unweighted N (Weighted %) | N/A | N/A | 5.5 (7.0) | 4.0 (5.6) | N/A | N/A |
| EC use every day  (5/5 days, %) | Weighted mean (SD) | N/A | 30 (47.4%) | N/A | 53 (84.1%) | N/A | 19 (48.4%) |
| Cotinine Serum  (ng/ml, mean) | Unweighted N (Weighted %) | 228.8 (149.0) | 227.9 (130.0) | 34.1 (106.2) | 248.0 (164.5) | 6.8 (38.6) | 98.1 (99.8) |
| N (%) | Unweighted N (Weighted %) | 349 (92.1%) | 71 (96.2%) | 67 (19.2%) | 58 (90.3%) | 23 (7.8%) | 36 (69.6%) |
| Albumin/Creatinine  N (%) | Weighted mean (SD) | 335 (93.0%) | 72 (93.1%) | 268 (92.7%) | 53 (85.5%) | 222 (93.8%) | 53 (99.7%) |
|  | URDACT 30-299 | 32 (5.8%) | 3 (6.9%) | 23 (6.1%) | 11 (14.1%) | 15 (5.7%) | 1 (0.3%) |
| **CKD (narrower definition)** | **URDACT ≥ 300** | **4 (1.1%)** | **0 (0.0%)** | **9 (1.2%)** | **1 (0.4%)** | **1 (0.5%)** | **0 (0.0%)** |
|  | URDACT **≥** 30 | **36 (7.0%)** | **3 (6.9%)** | **32 (7.3%)** | **12 (14.5%)** | **16 (6.2%)** | **1 (0.3%)** |
| eGFR (mL/min/1.73 m^2^, %) | eGFR < 60 | **4 (0.5%)** | **0 (0.0%)** | **16 (2.1%)** | **2 (1.5%)** | **0 (0.0%)** | **0 (0.0%)** |
| **CKD (broader definition)** | **eGFR < 60**  **or URDACT ≥ 30** | **37 (7.2%)** | **3 (6.9%)** | **37 (8.4%)** | **13 (15.1%)** | **16 (6.2%)** | **1 (0.3%)** |
| **CKD (narrower definition)** | **RR*** | ***Ref*** | **0.00** | ***Ref*** | **0.33** | ***Ref*** | **0.00** |
| **CKD (broader definition)** | **RR*** | ***Ref*** | **0.96** | ***Ref*** | **1.80** | ***Ref*** | **0.05** |

*RR calculated based on the prevalence of CKD among EC users divided by the prevalence of CKD among EC nonusers. No formal modeling analysis was conducted due to the limited sample size.

**Table S4** Matched, covariate-adjusted and population-weighted samples (matched by current, former, and never CC use)

| **Characteristic** | **Comment** | **Matched Case-Control** | |
| --- | --- | --- | --- |
|  |  | **EC Non-Use** | **EC Use** |
| N |  | 909 | 194 |
| Age (mean, SD) | Weighted mean (SD) | 35.8 (14.3) | 34.2 (14.4) |
| Male (%) | Unweighted N (Weighted %) | 550 (59.3%) | 114 (62.8%) |
| Non-Hispanic White (%) | Unweighted N (Weighted %) | 442 (70.1%) | 108 (75.9%) |
| Diabetes Dx or meds (%) | Unweighted N (Weighted %) | 68 (6.8%) | 12 (4.6%) |
| BMI ≥ 30 kg/m² (%) | Unweighted N (Weighted %) | 328 (37.0%) | 67 (32.2%) |
| BP, Systolic (mm Hg, mean/SD) | Weighted mean (SD) | 117.2 (13.6) | 118.3 (13.6) |
| BP ≥140 mmHg or taking antihypertensive medication(s) (%) | Unweighted N (Weighted %) | 180 (18.7%) | 35 (18.7%) |
| Current CC use (%) | Unweighted N (Weighted %) | 371 (42.6%) | 75 (36.4%) |
| Former CC use (%) | Unweighted N (Weighted %) | 300 (35.1%) | 65 (44.4%) |
| Never CC use (%) | Unweighted N (Weighted %) | 238 (22.4%) | 54 (19.3%) |
| Yrs. regular smoking  (mean, in current and former CC users) | Weighted mean (SD) | 19.8 (13.9) | 18.3 (14.0) |
| Current CPD  (mean, in current CC users) | Weighted mean (SD) | 12.7 (8.6) | 11.2 (9.8) |
| Time Quit CC  (yrs, mean, in former CC users) | Weighted mean (SD) | 5.5 (7.0) | 4.0 (5.6) |
| EC use every day (5/5 days, %) | Unweighted N (Weighted %) | N/A | 102 (63.9%) |
| Cotinine Serum (ng/ml, mean) | LBXCOT | 110.9 (155.4) | 211.8 (151.8) |
| Cotinine **≥** 10 ng/ml N (%) | LBXCOT **≥** 10 | 439 (47.7%) | 165 (88.5%) |
| Albumin/Creatinine N (%) | URDACT < 30 | 825 (93.1%) | 178 (91.0%) |
|  | URDACT 30-299 | 70 (5.9%) | 15 (8.8%) |
| **CKD (narrower definition)** | **URDACT ≥ 300** | **14 (1.0%)** | **1 (0.2%)** |
|  | URDACT **≥** 30 | 84 (6.9%) | 16 (9.0%) |
| eGFR (mL/min/1.73 m^2^, %) | eGFR < 60 | 20 (0.9%) | 2 (0.7%) |
| **CKD (broader definition)** | **eGFR < 60 or URDACT ≥ 30** | **90 (7.4%)** | **17 (9.3%)** |
| **CKD (narrower definition) *** | **RR*** | ***Ref*** | **0.17*** |
| **CKD (broader definition) *** | **RR*** | ***Ref*** | **1.26*** |

Note: estimates refer to unweighted N (weighted %) for categorical variables and the weighted mean (weighted SD) for continuous variables. *Calculated based on the prevalence of CKD among EC users divided by the prevalence of CKD among EC nonusers. No formal modeling analysis was conducted due to the limited sample size.
